# Sophisticated Prediction of Carotid-Plaque Vulnerability by Nanocluster Sensitized High-resolution Vessel-Wall-Imaging Profile in Rabbit Atherosclerotic Model

**DOI:** 10.1101/2023.03.09.23287077

**Authors:** Yan Gong, Menglin Wu, Dingwei Fu, Yu Guo, Xiudi Lu, Ying Zou, Xiang Zhang, Jinxia Zhu, Xianchang Zhang, Xue Li, Shuang Xia

**Affiliations:** Department of Radiology, Medical Imaging Institute of Tianjin, Tianjin First Central Hospital, School of Medicine, Nankai University, Tianjin, 300192, China; Department of Radiology, Second Hospital of Tianjin Medical University, Tianjin 300211, China; Department of Radiology, First Central Clinical College, Tianjin Medical University, Tianjin 300192, China; Department of Radiology, First Teaching Hospital of Tianjin University of Traditional Chinese Medicine, Tianjin, China National Clinical Research Center for Chinese Medicine Acupuncture and Moxibustion, Tianjin, China; Department of Radiology, Tianjin Nankai Hospital, Tianjin 300100, China; MR Collaboration, Siemens Healthcare Ltd., Beijing 100102, China

**Author notes:** Correspondencè Shuang Xia, Department of Radiology, Medical Imaging Institute of Tianjin, Tianjin First Central Hospital, School of Medicine, Nankai University, Tianjin, 300192, China, Xue Li, Department of Radiology, Second Hospital of Tianjin Medical University, Tianjin 300211, China, Email addresses. Statement of equal author contribution. Yan Gong, Menglin Wu and Dingwei Fu contributed equally to this study Shuang Xia and Xue Li are co-correspondences.

**Keywords:** Atherosclerosis, plaque, high-resolution magnetic resonance vessel wall imaging, histogram, macrophage-target nanoparticle

## Abstract

**OBJECTIVE:** To innovatively developed a macrophage-target nanoparticle based contrast-enhanced high-resolution magnetic resonance vessel wall imaging (HR-VWI) strategy to characterize the plaques’ vulnerable features on rabbits.

**BACKGROUND:** Lacking of sensitive and specific image-marker of HR-VWI leads this technique depending upon the plaque morphological characteristics. Nanoparticle-based contrast agents modified with targeting ligands allow amplifying MR signals of the interested components. The key to successful translation is the requirement that conducting studies in larger animals to provide reasonable diagnostic readouts.

**METHODS:** The HR-VWI enhanced with macrophage-targeted PP1-Au@GSH@Gd (GdMG) nanoclusters (NCs) and the conventional Gadovist were utilized for the plaque vulnerability evaluation by a systematic histogram analysis in atherosclerosis (AS) rabbit model.

**RESULTS:** Due to the compelling targeting capacity of GdMG NCs to foamy macrophages, the contrast-to-noise ratio (CNR) from pre-injection baseline dramatically raised from 6.50 to 36.91 (*p* < 0.001), with an increment of 1.39-fold higher than that of the Gadovist approach. Spearman’ s correlation test confirmed that the coefficient of variation (CV) derived from the histogram analysis based on GdMG NCs HR-VWI was indeed positively linearly correlated with pathology vulnerability index (VI_P_) significantly (*p* < 0.05) with adjusted R^2^ = 0.775. Finally, mathematic formulas with histogram-derived parameters as variables were fitted to quantitatively calculate the histogram vulnerability index (VI_H_) with the strength of the adjusted R^2^ = 0.952 (*p* < 0.001), and Area under the curve (AUC) of 0.875 (*p* < 0.001) to realize the *in vivo* and quantitative calculation of the plaque vulnerability.

**CONCLUSION:** Profiting from the splendid inflammation targeted capacity and excellent MRI performance of GdMG NCs, as well as the highly quantitative characteristics of histogram analysis, we disclosed that our established imaging protocol was able to identify the plaques’ vulnerability index that were comparable to pathological examinations in both retrospective and prospective experiments.

## 1. INTRODUCTION

Ischemic stroke that attributed to thromboembolism caused by carotid atherosclerotic plaque has represented a massive public health issue^1–3^, with an estimated 6.5% increase in mortality from 2015 to 2019^4^. These sudden ischemic strokes usually contribute to unheralded rupture of high-risk atherosclerotic plaques. In this context, identification of plaque features which pose a high risk of rupture in a noninvasive manner is of paramount important for preventing acute vascular events. Among all the current imaging techniques, contrast-enhanced high-resolution magnetic resonance vessel wall imaging (HR-VWI) has been recognized as a promising imaging technique to track the arterial atherosclerosis by depicting the plaque presence, size and morphology with submillimetre resolution. A useful work in this term by our group has demonstrated that contrast-enhanced HR-VWI is a compatible standard to digital subtraction angiography (DSA) on analyzing plaque morphology stenosis, benefiting from its efficient suppression on surrounding fluid signals and high temporospatial resolution^5^. Despite well appreciated advantages, the inherent limitations of HR-VWI, especially lack of sensitive and specific MRI contrast agents, lead this imaging technique being largely dependent on the morphological characteristics of the plaques when stratifying atherosclerotic risk, severely hampering its application in preventing acute vascular events. In some cases, the risk plaques appear completely normal without any prior symptoms, and the changes of indicative biomarkers such as intraplaque hemorrhage (IPH), lipid-rich necrotic core (LRNC), thin fiber cap are prerequisitely required for confirmation of the plaques’ diagnosis^6–8^.

Tremendous efforts have been devoted to the development of MRI contrast agents to diagnose atherosclerosis beyond morphological characteristics or physiological function changes. Particularly, nanoparticle-based contrast agents with high surface-to-volume ratios allow the surface layers to be modified with targeting ligands (e.g., antibodies, proteins, peptides), for targeted local amplifying MR signals of the interested components^9–11^. For example, more recently, our groups reported the peptide functionalized nanoparticles that can specifically target inflammatory macrophages in plaques for quantitatively characterizing the activity of foamy burden at the molecular level in ApoE^−/−^ mice^12^, providing a chance for the utilization of nanoparticle-based contrast agents to predict the carotid plaque vulnerability^13^. Although the MR signals generated by the nanoparticle-based contrast agents are associated with the vulnerability of plaques, it still remains daunting gap between the pre-clinical researches and large-scale clinical applications. One of those challenges is that overwhelming majority of pre-clinical works is merely limited to proof-of-concept mouse studies. However, the key to successful translation is the need to conduct studies in larger animals to provide reasonable diagnostic readouts that differ from mouse studies. Another challenge is the interpretation of the MR signal patterns still highly relies on the radiologists’ expertise and experience, which may reduce the reproducibility or consistency of diagnostic results, and thereby impairing the long-term monitoring of atherosclerosis progression. Concurrently, a paradigm shift in this field has gained momentum as new imaging strategies to provide useful experience for the clinically translation.

To address this need, we report a nanoparticle (Gd-modified macrophage-target gold nanoclusters, GdMG NCs) based imaging strategy that surmounts the aforementioned hurdles to precisely predict the carotid plaque vulnerability on the medium-sized animal model (a rabbit atherosclerotic model). Capitalizing on established work^12^, we firstly confirmed the HR-VWI performance of GdMG NCs *in vitro* and *in vivo*. Next, we innovatively developed a systematic histogram-based imaging protocol to characterize the plaques’ vulnerable features through establishing the relationships between the sensitized HR-VWI histogram profile and the histological vulnerability index. As anticipated, the *in vivo* performance of GdMG NCs suggested that GdMG NCs could promote HR-VWI recognition of AS plaques, evaluating plaque progression and simultaneously identifying the plaques’ vulnerability index in a noninvasive manner. Profiting from the splendid inflammation targeted capacity and excellent MRI performance of GdMG NCs, as well as the highly quantitative characteristics of histogram analysis, we disclosed that our established imaging protocol was able to identify the plaques’ vulnerability index that were comparable to pathological examinations in both retrospective and prospective experiments.

## 2. METHODS

### 2.1 SYNTHESIS OF GDMG NCS

The synthesis of the PP1-Au@GSH@Gd NCs (GdMG NCs) followed a typical pattern. The synthesis of GdMG NCs and *in vitro* parallel imaging experiment is analytically described in Supplemental Material (*Synthesis of PP1-Au@GSH@Gd NCs*).

### 2.2 ANIMAL MODEL OF ATHEROSCLEROTIC PLAQUE

The plaque formation was induced by balloon injury on the rabbits left common carotid artery (LCCA) and followed with continuously high-fat diet (HFD). The atherogenic rabbits were intramuscularly anesthetized with ketamine (35 mg/kg), acepromazine (0.75 mg/kg), and xylazine (5 mg/kg). During the procedure, anesthesia was maintained with isoflurane inhalation. Rabbits were euthanized with an overdose of sodium pentobarbital (160 mg/kg i.p.). The feeding protocol and surgery procedure was adequately described in Supplemental Material (*Animal model of atherosclerotic plaque*).

### 2.3 IN VIVO HR-VWI AND ANALYSIS

The quantitative imaging measurements on the source images and the reconstructed images for the vessel wall area (WA) (**Figure 2G**), and plaque area (PA) (**Figure 2G**) are given. The contrast-to-noise ratios (CNR) and the contrast index (CI) were measured to reflect the enhancement ability and nanoclusters deposition in the plaque region. The quantitative histogram analysis of HR-VWI data was performed by segmenting the plaque and using the freehand selection tool in the ImageJ software on HR-VWI (**Figure 4A**).^14^ The detailed HR-VWI examination criteria, applied sequence protocol and analysis criteria are described in the Supplemental Material (*In vivo HR-VWI and analysis, Standard workflow of image interpretation* and **S. Table 1**).

**Figure 1.**
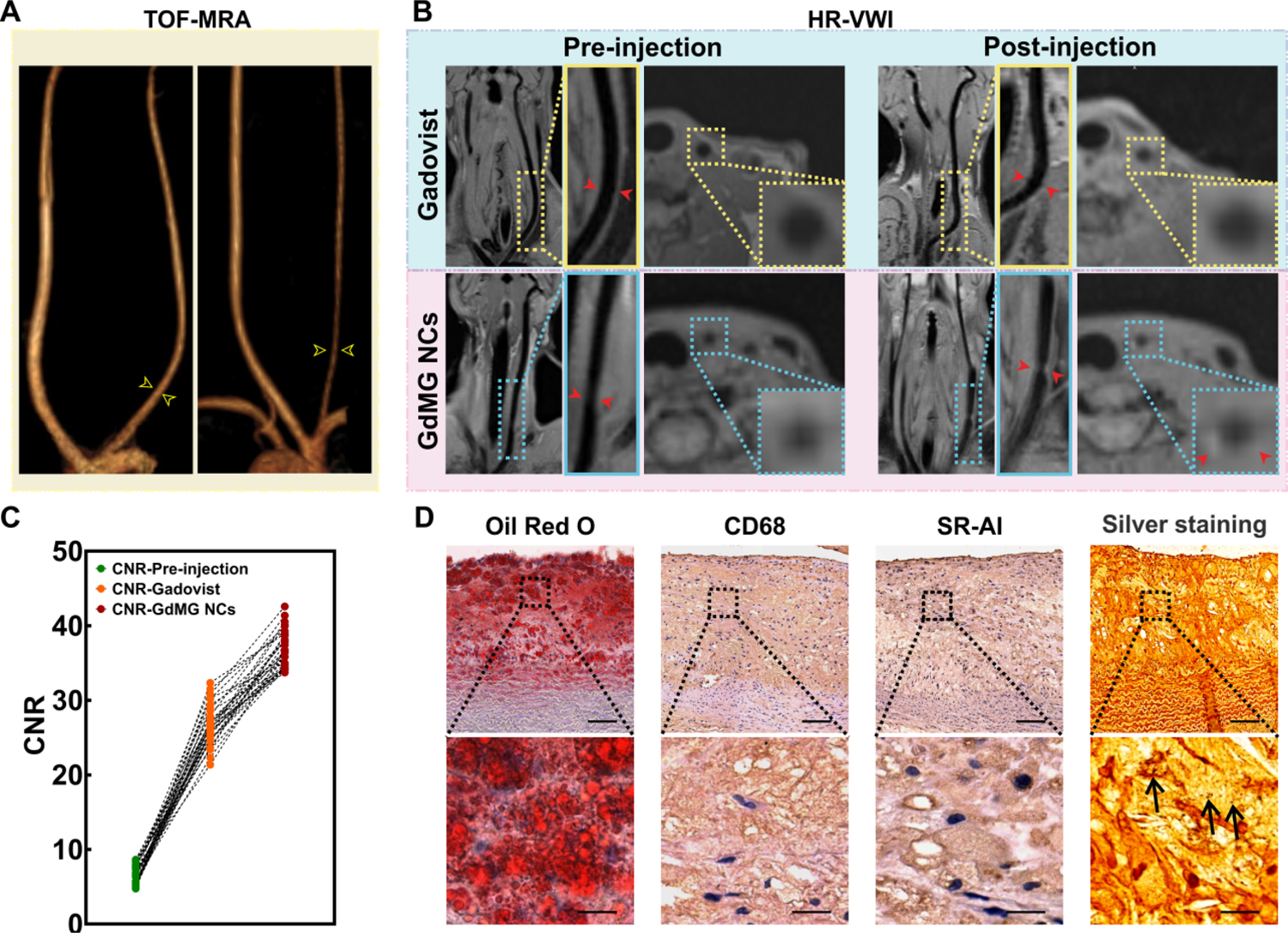
Construction of Atherosclerotic Rabbit Model. The experimental timeline (A) includes a one-week regular diet, a one-week atherogenic diet before the injury, balloon injury, and atherogenic feeding before euthanasia. The blood lipid disorder (B) (TC: total cholesterol; TG: triglycerides; HDL-C: high-density lipoprotein-cholesterol; LDL-C: low-density lipoprotein) were induced by high-fat diet (HFD) and the body weight (C) keep increasing during the experiment session. Gross lesions of the aorta and carotid artery, and stained by Oil red O (visualized as a red area on face) (D) shows the atherosclerosis change. Serial paraffin cross-sections of the carotid artery were stained with Oil red O (E) revealing the growth of plaque lipid continent over time.

**Figure 2.**
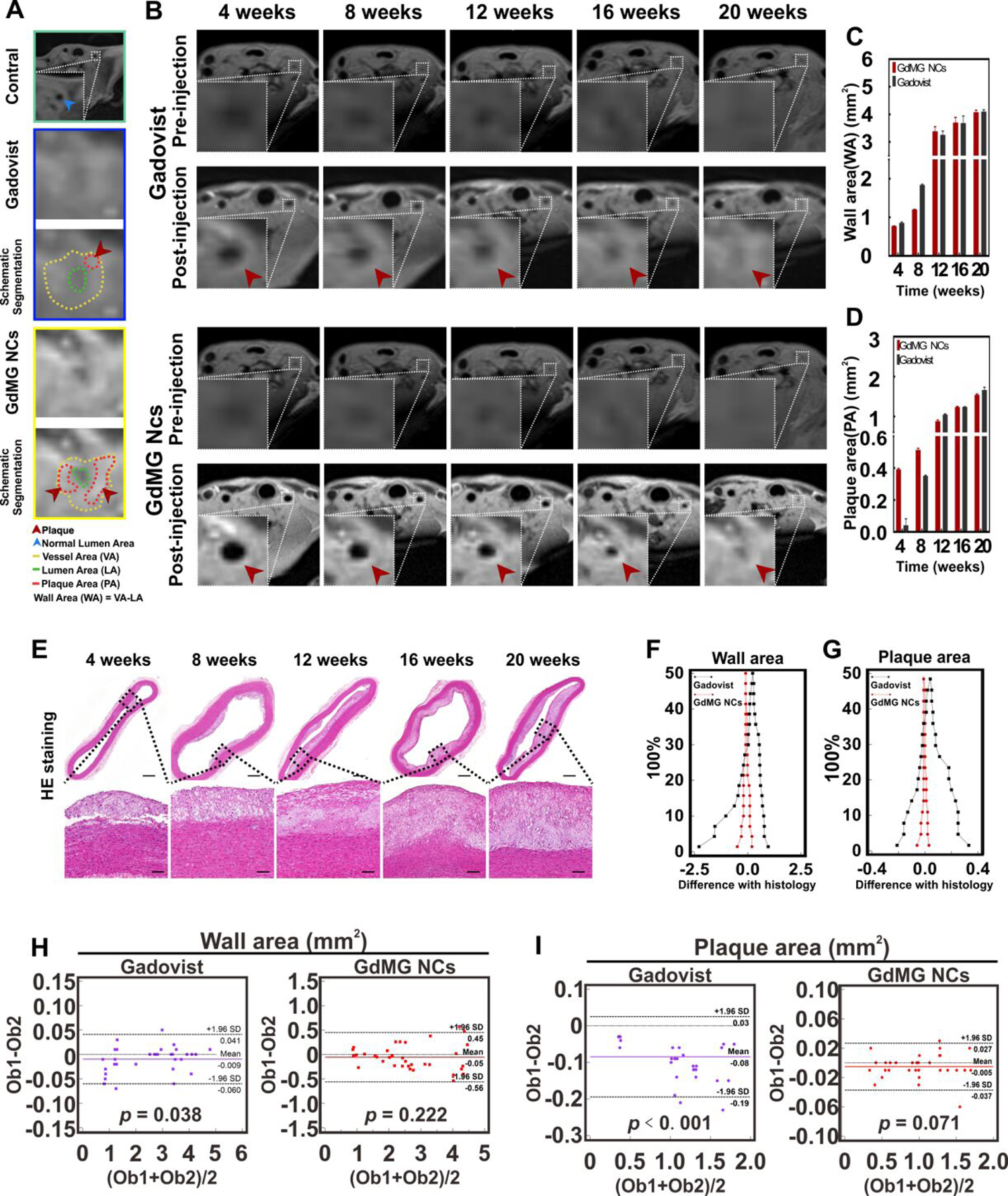
In vivo target ability of GdMG NCs in atherosclerotic rabbits. The vessel wall images of atherosclerotic rabbit carotid artery before and after the injection of contrast agent (highlighted within a yellow box). (A) The traditional TOF-MRA clarifies the vessel diameter to measure the severity of atherosclerosis (yellow arrowhead). (B) Contrast-enhanced HR-VWI, from which GdMG NCs were able to identify a higher eccentric plaque signal on the axis section image (red arrowhead), while Gadovist only showed the vessel wall thickening with circumferential enhancement (yellow dotted box). The contrast to noise ratio (CNR) (C) GdMG NCs was significantly higher than that of Gadovist (*p* < 0.05).The histology assessment of the stenotic lesions was conducted for the validation of the GdMG NCs targeting capability to macrophages in plaque (D). Abundant lipid deposits were shown by Oil red O staining, confirming that the enhanced areas were definite plaque sites; over-expression of CD68 and SR-AI were detected by IHC staining, indicating the foamy macrophages tremendously enriched in plaque area (black arrow); the extensive presence of GdMG NCs deposition in plaque was validated by silver-staining (black arrow), which further providing strong shreds of evidence to the compelling targeting capacity of GdMG NCs to atherosclerotic plaques.

### 2.5 EX VIVO ANALYSES

The histology staining for plaque tissue and the analysis criteria of pathology vulnerability index (VI_P_) for differentiating vulnerable plaques was meticulously described in the Supplemental Material (*Ex vivo analyses*).

The characterization and stability study of GdMG NCs in combination with *in vivo* security evaluation was also described in Supplemental Material (*Characterization of PDA/Gd/Cu* and **S. Figure 1**), as has been reported in the previous works in the literature.^15–17^

## 3. STATISTICAL ANALYSIS

The continuous variables were reported as appropriate. The normality was assessed formally by using the Shapiro-Wilk test. The advanced statistical methods were described in Supplemental Material (*Statistical Analysis*).

## 4. RESULTS

### 4.1 CONSTRUCTION OF THE ATHEROSCLEROTIC RABBIT MODEL

The successful construction of the atherosclerotic rabbit model can be observed from **Figure 1** and were described in Supplemental Material (*Supplemental Results*).

### 4.2 INVESTIGATION OF THE HR-VWI PERFORMANCE OF GDMG NCS

In this work, for targeting the imaging foamy macrophage in plaque, the PP1-Au@GSH@Gd NCs (GdMG NCs) was constructed by in situ reducing the Au atom into the zwitterionic glutathione (GSH) shell with both Gd–DTPAA and PP1 peptide modification, which yielded uniform size and a superior degree of dispersion. The characteristics of the prepared GdMG NCs were described in (supplemental **S. Figure 1**). Then, the imaging property of GdMG NCs on HR-VWI was investigated *in vitro* and *in vivo* (supplemental **S. Figure. 2**) .From these results, strong shreds of evidence were provided to verify the GdMG NCs could serve as an ideal contrast agent for HR-VWI.

Encouraged by the superb performance of *in vitro* HR-VWI contrast capacity, the *in viv*o foamy macrophage targeting and the selectivity efficacy of GdMG NCs on HR-VWI in atherosclerotic rabbit models were next comprehensively quantified in comparison with Gadovist (**Figure 2**). The results proved that GdMG NCs providing more definite morphologic indices to identify the manifestation of culprit plaque-induced stenosis in LCCAs compared with the Gadovist approach (detailed in *Supplemental Results*, **S. Figure 3 and S. Figure 4**). Consequently, from the Oil Red O, IHC staining, and silver-staining of the LCCAs, significant lipid deposits and abundant foamy macrophages enrichment were verified in the HR-VWI-highlighted regions induced by the GdMG NCs aggregation, providing strong pieces of evidence for the compelling targeting capacity of GdMG NCs to foamy macrophages in atherosclerotic plaques.

**Figure 3.**
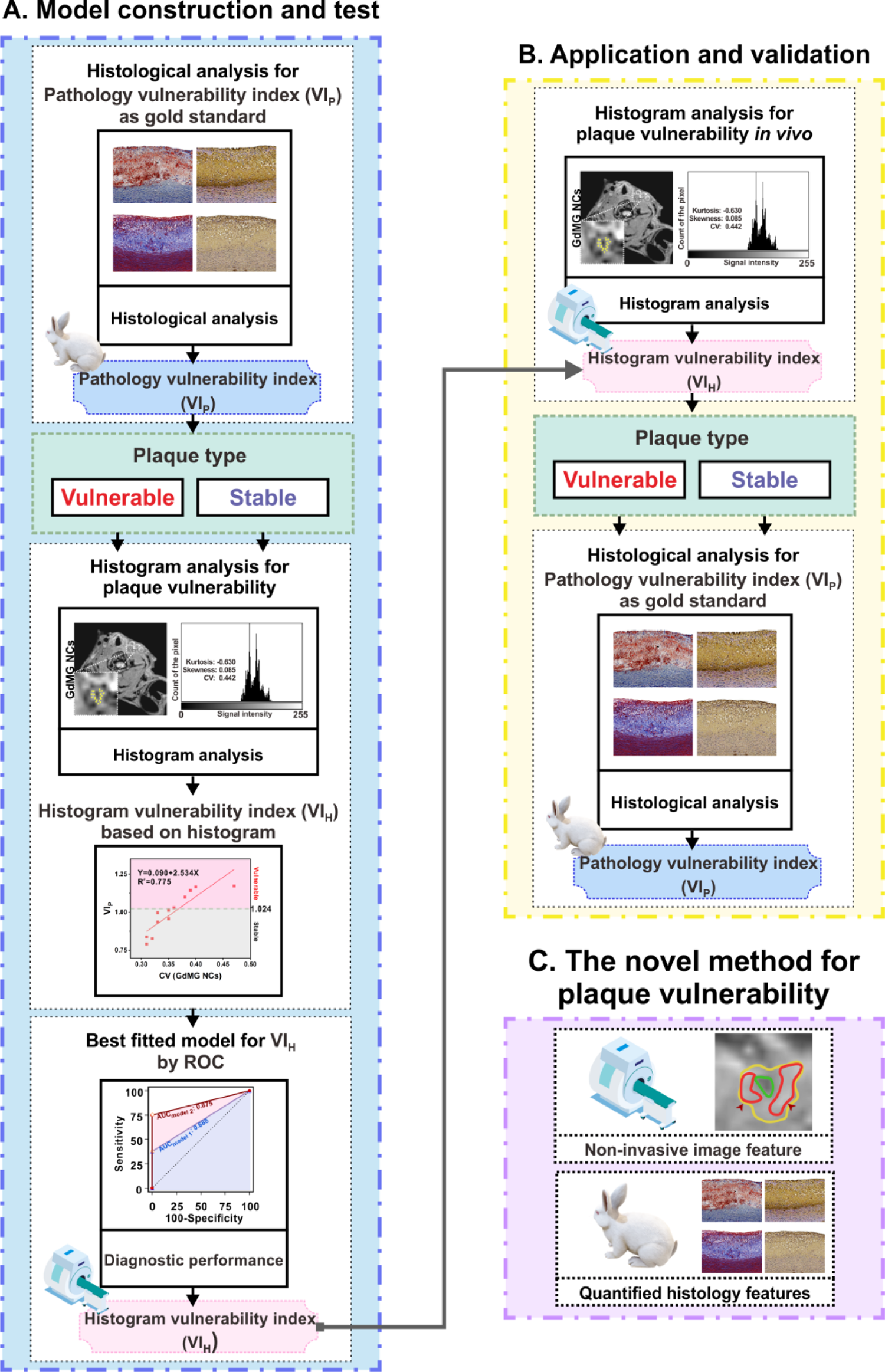
Dynamic changes of wall area (WA) and plaque area (PA) of atherosclerotic rabbit’s carotid artery by contrast-enhanced HR-VWI with GdMG NCs. (A) Schematic segmentation of HR-VWI. The control case showed a cross-section image of normal rabbit LCCA with smooth, round and non-stenotic lumen inner-surface. As can be ascertained, the crescent-shaped hyperintense signal (representing PA, red line) enhanced by the presence of GdMG NCs was significantly higher than Gadovist, rendering the clearer plaque delineation. Besides, the eccentric iso- and slightly hyperintense signal representing WA (area between yellow and green circle) could also be easily delineated by the incorporated GdMG NCs with sharp demarcation. While after the Gadovist administration, WA presented a heterogeneous annular enhancement region with blurred boundaries, in which the existence of a relatively ambiguous contrast-enhanced spot (recorded as PA) could be barely outlined manually. (B) The cross-section images represent a crescent-shaped hyperintense signal (representing PA) enhanced by GdMG NCs was significantly higher than Gadovist (red arrowhead). The dynamic progression of plaque WA (C) and PA (D) was illustrated according to the contrast-enhanced HR-VWI with Gadovist and GdMG NCs. H&E-staining sections (E) determine the true histological PA and WA at different plaque stages. The percentile for the rated difference between contrast-enhanced HR-VWI to pathology is shown in mountain plots (F, G). The GdMG NCs curve (red dot-dash-line) is steeply centered around zero, indicating a small divergence from histology than Gadovist (black dot-dash-line). Bland-Altman plot (H) shows the comparison of the inter-observer agreement for contrast-enhanced HR-VWI evaluation. GdMG NCs could offer a more satisfying diagnostic agreement than Gadovist, since GdMG NCs have less inter-observer difference (red dots, *p* > 0.05) than Gadovist (purple dots, *p* < 0.05). \**p* < 0.05, n = 6 biologically independent animals in different plaque stages. \**p* < 0.05, Bonferroni adjustment for multiple comparisons, n = 6 biologically independent animals in each time point. The data are presented as mean ± SEM. RM ANOVA with Bonferroni’s post hoc test.

### 4.3 THE REPRODUCIBILITY AND RELIABILITY OF PLAQUE EVALUATION BY GDMG NCS ENHANCED HR-VWI

Herein, the dynamic changes of the wall area (WA) and plaque area (PA) were also used to quantify the collected imaging patterns (**Figure 3A**). For the progression of WA (**Figure 3C**) and PA (**Figure 3D**) determined by the two different contrast agents, repeated measures analysis of variance (RM ANOVA) was performed. By referring to the gold standard in the histology of the H&E staining (supplemental **S. Figure 7**), the curve of GdMG NCs was more steeply centered to the base axis (x = 0), indicating better accordance with the histological analysis compared to Gadovist (*p <* 0.05) approach for the assessment of WA (**Figure 3F**) and PA (**Figure 3G**) (supplemental material S. Table 3).^18^ The detailed results Intra-observer, and inter-observer reproducibility of both GdMG NCs and Gadovist in the quantification of WA and PA were described in Supplemental Material (*Supplemental Results*).

### 4.4 IN VIVO EFFECTS OF GDMG NCS ON PLAQUE VULNERABILITY PREDICTION

After revealing the superior competence of GdMG NCs in measuring the morphological assessment for plaque, plaque vulnerability prediction studies were further conducted based on the contrast-enhanced HR-VWI. **Figure 4** presents a graphical flowchart of the designed method for the quantitative characterization of plaque vulnerability by histogram analysis based on GdGM NCs enhanced HR-VWI *in vivo*. The pathology vulnerability index (VI_P_), which can be defined as the ratio of the destabilizing components (macrophage plus lipid) to the stabilizing components (smooth muscle cell plus collagen), was used as the gold standard for quantifying plaque vulnerability, with stable (VI_P_ < 1.024) and vulnerable (VI_P_ ≥ 1.024) groups^19^. VI_P_ emphasized the destabilized role of macrophage in plaque vulnerability, which additionally reflected the plaque heterogeneity comprehensively. According to these comprehensive criteria, histological-defined plaques were divided into vulnerable and stable groups to retrospectively analyze their histogram characteristics on GdMG NCs enhanced HR-VWI. As is illustrated in **Figure 5A**, the shapes of the histogram by GdMG NCs dramatically varied in different plaque types. To quantify the histogram features in the different plaque types, the kurtosis, skewness, and CV were utilized to clarify the correlation between the signal intensity distribution and VI_P_. The histological classification for plaque vulnerability that corresponds to the image-colocalization in **Figure 5B** may provide clues to the histogram features presented by vulnerable plaques. As can be observed from **Figure 5C**, CV derived from GdMG NCs of vulnerable plaque was significantly higher than that of the stable plaque (*p* = 0.003), indicating the existence of a more dispersed signal distribution representing the heterogeneous enhancement (supplemental **S. Table 2**), which can be explained by the above-mentioned enhancement mechanism related to the uneven distribution of macrophages. Furthermore, from the Spearman’s correlation test, it was confirmed that the CV was indeed positively linearly correlated with VI_P_ significantly (*p* < 0.05) with adjusted R^2^ = 0.775 (**Figure 5D**, supplemental **S. Table 7**), implying a positive correlation between CV and macrophage content as well. Comparatively, other indicators (kurtosis and skewness) turned out to have no significant difference for dichotomous outcomes (stable vs vulnerable) derived from GdMG NCs (supplemental **S. Figure 9**). Nonetheless, none of the histogram parameters derived from the Gadovist method showed distinctions between stable and vulnerable plaques (*p* > 0.05). Simultaneously, by considering the VI_P_ was time-dependent (**Figure 5E**) (*p* < 0.05), both the GdMG NCs derived histogram parameters and HFD-inducing period (Week) were introduced into the multiple linear regression analysis to establish the non-invasive histogram vulnerability index (VI_H_) for reasonable prediction. Meanwhile, from the histological classification results with VI_P_ for the overall assessment of plaque composition, an increasing proportion of vulnerable plaques during long-term HFD intervention was demonstrated (supplemental **S. Figure 10**). Finally, two formulas were fitted as follow: (model 1) VI_H_ = 0.366 + 0.012Week + 1.361CV, and (model 2) VI_H_ = 0.226 + 0.005Week + 2.459CV - 0.019kurtosis. In model 1, CV and Week contributed as variables to calculate VI_H_ with adjusted R^2^ = 0.897 (*p* < 0.001). Notably, the strength of model 2 was stronger elevated with CV, kurtosis, and Week as the variables with different coefficients for VI_H_ with adjusted R^2^ = 0.952 (*p* < 0.001) (supplemental **S. Table 8**). Based on the above-mentioned results, *in vivo* and quantitative calculation for plaque vulnerability was realized by using histogram-defined parameters (histogram vulnerability index, VI_H_) derived from GdMG NCs enhanced HR-VWI.

**Figure 4.**
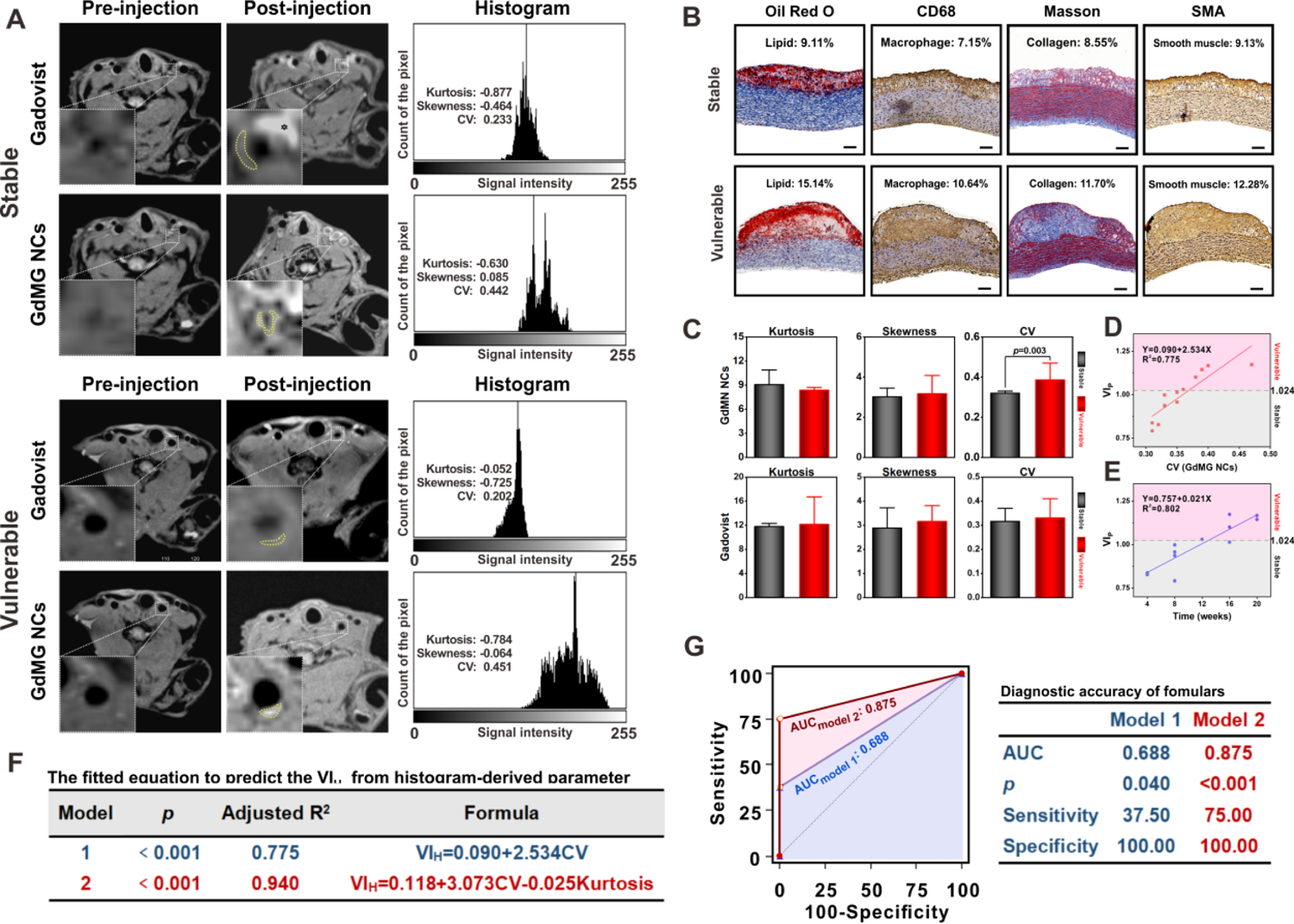
Flowchart of the *in vivo*, non-invasive and quantitative characterization of plaque vulnerability and the final performance evaluation in the work.

**Figure 5.**
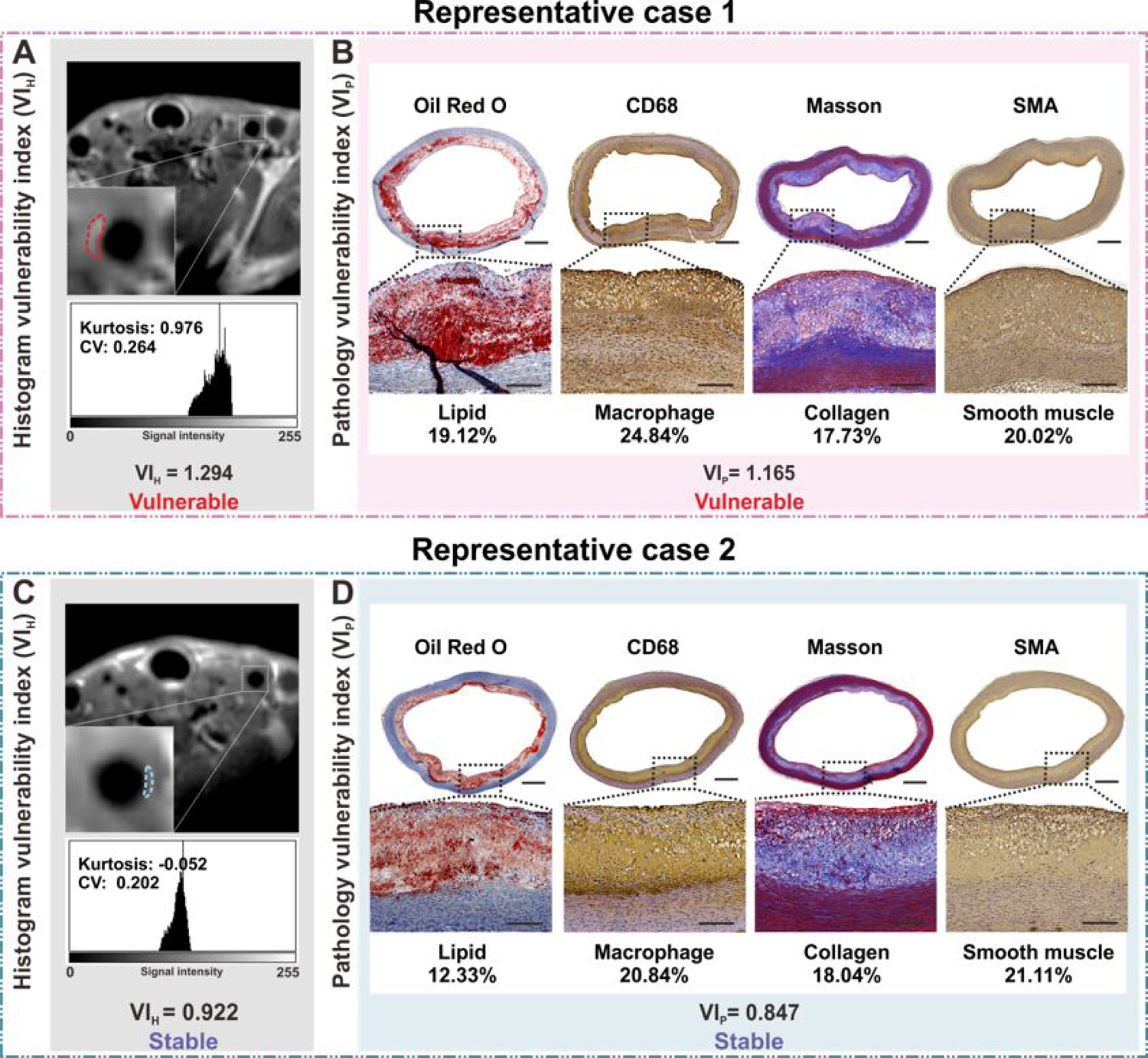
Non-invasive Plaque vulnerability prediction by GdMG NCs enhanced HR-VWI images and histogram analysis. The histogram analysis (A) of plaque (yellow dot-line) according to contrast-enhanced HR-VWI with Gadovist and GdMG NCs. The signal distribution features are described by skewness (asymmetrical shape), and kurtosis (sharpness), and CV describes the dispersion of signal intensity. The serial co-registered histology tests (B) are quantified and analyzed to calculate the pathology vulnerability index (VI_P_) for distinguishing vulnerable plaques (VI_P_≧1.024) from stable plaques (VI_P_≧1.024). Box plots (C) showed the difference in the histogram parameters and signal intensity features between stable (n = 17) and vulnerable plaque (n = 13), evaluated on contrast-enhanced HR-VWI (\**p* < 0.05). 3D The scatter plots (D) describe the relationship among kurtosis, skewness and CV, with the color mapping representing the VI_P_ value and the scatter diameter representing the plaque area. The fitting formula based on the multiple linear regression model (E) offers a theory for calculating the VI_P_ from the contrast-enhanced HR-VWI *in vivo*. The fitting formula from the GdMG NCs (model 2) committed a better diagnosis prediction for vulnerable plaque. Besides, CV showed a significant correlation to VI_P_ (*p* < 0.05).

Consequently, the diagnostic accuracy and prediction performance of VI_H_, which was calculated from the fitting formulas (models 1 and 2), was further prospectively investigated in distinguishing vulnerable plaque *in vivo* with histology validation. For this reason, twelve atherosclerosis rabbits were randomly chosen covering the whole HFD-inducing session and were imaged by GdMG NCs enhanced HR-VWI for the VI_H_ evaluation. The histogram-derived parameters based on GdMG NCs enhanced HR-VWI were next introduced to the fitting formulas of the VI_H_ prediction. Then, the efficiency of the prediction results was confirmed by the histological proof (VI_P_) with a total number of 8 vulnerable plaques and 4 stable plaques. Supplemental S. Table 9 depicts the sum of 3 (3/8) lesions with VI_H_ ≥ 1.024 that were precisely assigned to vulnerable plaques by using formula model 1, whereas model 2 detected 6 (6/8) vulnerable ones. On the contrary, 5 (5/8) vulnerable plaques were misestimated as stable lesions by using the formula model 1, and only 2 (2/8) were misestimated by model 2. Meanwhile, 4 stable plaques were all accurately recognized by both formulas. The above-mentioned results indicated that formula model 2 had a higher discriminative power in identifying the high-risk vulnerable plaques. The ROC analysis (**Figure 5G**) was also conducted to examine the diagnostic performance of the two models in determining the vulnerability of each lesion against the reference standard (VI_P_). What’s more, Model 2-defined improved the diagnostic accuracy with an AUC value of 0.875 (*p* < 0.001), which was 1.27-fold higher than model 1 (AUC = 0.688, *p* = 0.040). Additionally, as an image-mediated diagnostic protocol, both sensitivity and specificity for potential risk prediction should be aimed to improve. The table in **Figure 5G** illustrated the significantly elevated diagnostic sensitivity of model 2 with respect to model 1 (75.0% vs 37.5%), maintaining the perfect diagnostic specificity (100% vs 100%) as well.

### 4.5 DIAGNOSIS AND PREDICTION OF VULNERABLE PLAQUE VIA NANO-ENHANCED IMAGING VULNERABILITY INDEX

The representative cases are shown in **Figure 6**. The VI_H_ results of case 1 (**Figure 6A**) by model 2 deemed it a vulnerable lesion (VI_H_ = 1.294). From the histological analysis (**Figure 6B**), a vulnerable plaque (VI_P_ = 1.165) was confirmed, which was in accordance with model 2. Case 2 (**Figure 6C**) revealed also the stable lesion consistently diagnosed by model 2 (VI_H_ = 0.992), which was further acknowledged by histology (VI_P_ = 0.847) (**Figure 6D**). From the above-mentioned results, it was demonstrated that the utilizing value is a superb predictive tool for determining plaque vulnerability by using the formula model 2 extracted from GdMG NCs enhanced HR-VWI, both non-invasively and quantitatively. The histogram-defined signal dispersion, CV, is a strong predictor for the differentiation of the lesion type for both MCA and BA. Compared with CV, the combination of other parameters including kurtosis, and time did improve the diagnostic power significantly.

**Figure 6.**
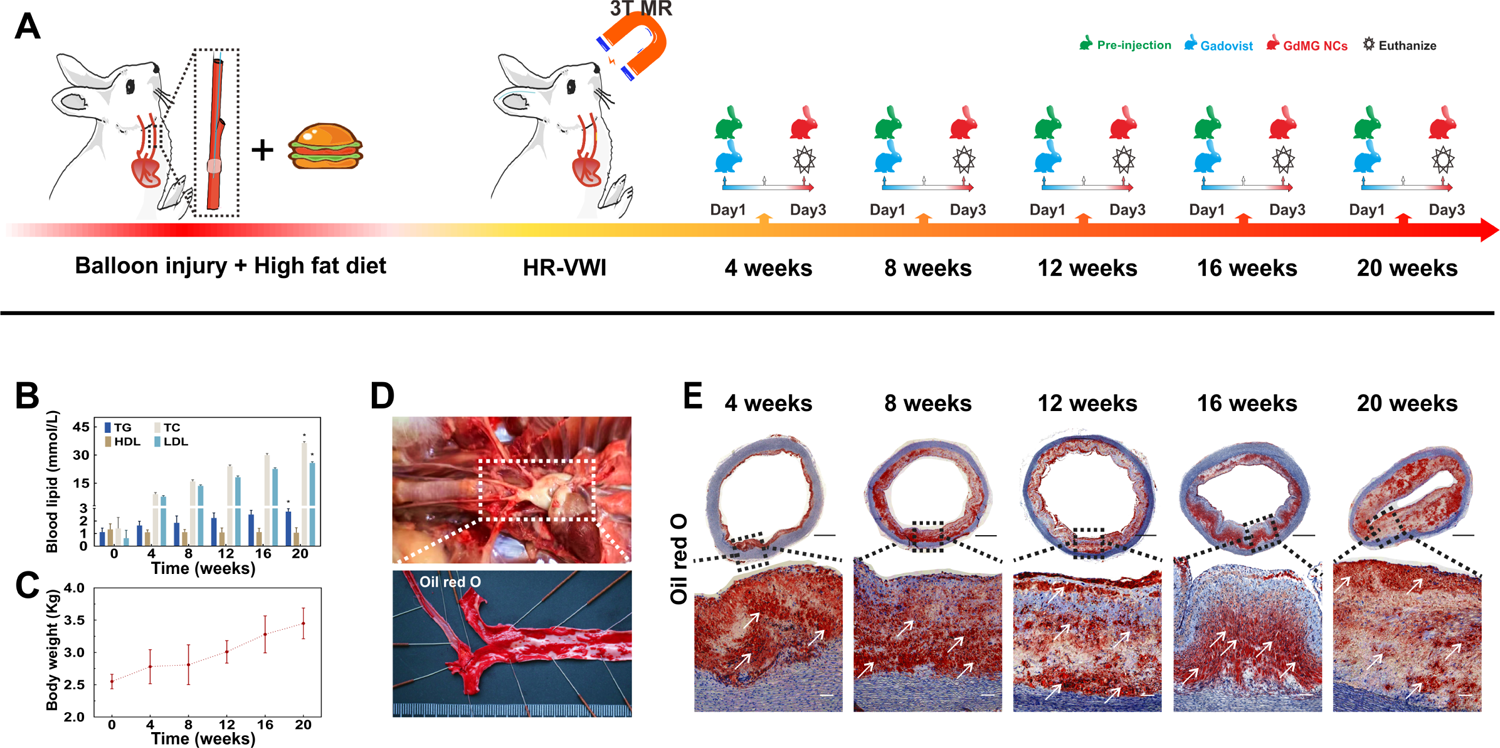
The validation of the diagnostic value of the GdMG NCs enhanced HR-VWI. (A) Histogram vulnerability index (VI_H_) was calculated based on contrast-enhanced HR-VWI with Gadovist and GdMG NCs. (B) Pathology vulnerability index (VI_P_) calculated as the gold criteria for plaque vulnerability. (C) Receiver operating characteristic curve for evaluating our prediction models. The area under curve (AUC) represents the proposed fitting formula, especially model 2 has a perfect diagnostic performance.

### 4.6 BIOCOMPATIBILITY OF GDMG NCS

The Biocompatibility of GdMG NCs in Supplemental Material (*Biocompatibility of GdMG NCs* and **S. Figure 11**) proved that it had no remarkable side impact on tissues, further confirming its prominent biocompatibility, which might carve out the way for its promising future biomedical applications.

## 5. DISCUSSION

A remarkable gain for the accurate assessment of plaque vulnerability was demonstrated by this innovatively designed work through macrophage-targeted enhanced HR-VWI-based histogram analysis. The diagnostic and predictive value of the proposed method in quantitatively distinguishing vulnerable plaques in perfect accordance with the respective histology criteria (pathology vulnerability index) was highlighted. Furthermore, the combination of HR-VWI, macrophage-targeted contrast agent, and histogram analysis can offer a direct mechanistic link to improve the diagnostic reliability objectively, thereby facilitating the clinical translation.

The advantages of HR-VWI and the necessity of the macrophage-targeting contrast agents were discussed in Supplemental Material (*Supplemental discussion*)

It is a critical-but-challenging task for the current image-evaluation method to provide objective and biologic-accordance information that helps in both clinical diagnosis and management. Growing pieces of evidence have demonstrated the morphological, pathological, and functional properties of the various tissue types that are associated with a high likelihood of plaque rapid progression and rupture^20^. More specifically, a large LRNC infiltrated by macrophages was prone to rupture, while, on the contrary, collagen, and plaque smooth muscle cell help stabilize the plaque structure. In general, the pathology vulnerability index (VI_P_) is considered an important index to screen vulnerable plaques the higher VI_P_ represents more unstable plaques, whereas a VI_P_ larger than 1.024 was proved to be vulnerable. “Vulnerable plaque” can be viewed as a function of its composition than size. Meanwhile, the vulnerable plaque relies on image interpretation was diagnosed in previous studies, which may overlook or overestimate some morphological information (lumen stenosis, plaque enhancement, and IPH) due to personal diagnostic experience and understanding of the MR physics in radiology.^6,21^ Hence, to better access the plaque vulnerability, it is vital to emphasize the tissue-specific image properties of the plaque heterogeneity by image interpretation. Inspired by SHI Z. etc., who reported that histogram analysis based on conventional T_1_WI image can contribute to differentiating the intracranial plaque type, we expanded their conclusion and finally reached an exciting advance in quantified plaque vulnerability with a statistical method. From the provided statistical analysis, it can be argued that CV has a positive correlation to plaque type, which was also consistent with the literature.^22^ The underlying reason for vulnerable plaque to exhibit such high CV may be due to the various components in advanced lesions, i.e. necrosis increased due to the impaired ability of macrophages to efferocytose apoptotic cells, and macrophages in advanced lesions also secrete matrix metalloproteinases contributing to fibrous cap thinning and plaque rupture.^23, 24^ In addition, GdMG NCs can also generate HR-VWI signal increment when internalized by macrophages. Thus, the ratio of the foamy macrophage in plaque can impact the kurtosis (reflecting the most range of signal intensity)^25, 26^. Skewness also reflects the uneven plaque component distribution by the existence of an asymmetrical shape. For example, the distribution of vascular smooth muscle cell (VSMC) and micro-calcification, both of which may increase the plaque rupture risk.^27^ Furthermore, the histogram diagram can fully reflect the plaque heterogeneousity since the different components have distinguishing signal patterns due to their inherent relaxation rate within the ROI. To the best of our knowledge, our designed experiment is the pioneering workflow to quantitate the plaque vulnerability via histogram based on the macrophage-targeted plaque HR-VWI *in vivo*, which had a satisfactory diagnostic performance with both optimized specificity and sensitivity. Moreover, the proposed analysis processing is generated in Radiant and ImageJ software packages, both of which are in general use and easily available, hence making the introduced method applicable and adjustable for other studies in plaque vulnerability and clinical applications for risk management.

Although the high sensitivity of the HR-VWI-based molecular image method permits the detection of subtle morphological and physiological changes of AS plaque with small sample sizes, this animal experiment still suffers from several limitations including essential differences in disease progression from humans, lack of external validation for clinical management, and long duration to assess disease progression.^28–30^ Here, the concept of triangulation by image-evaluating, histology validation, and statistical analysis were applied to increase confidence that an objective exact method was correctly conducted.

## 6. CONCLUSIONS

The proposed integrative analysis provided an overall picture of the non-invasive nano-image method for quantifying plaque vulnerability and substantially increasing the relevance of the results for clinical reference. The use of GdMG NCs as macrophage-targeted HR-VWI is considered a promising diagnosis avenue for many inflammation-related diseases. Additionally, a deep understanding between the quantified histogram parameters and plaque vulnerability will potentially improve our ability to harness the mechanism for developing novel image strategies against cardiovascular disease and promote the prediction before adverse clinical events. Although this is a proof-of-concept that was only tested in rabbits, it lays a strong foundation for the bench-to-bedside translation of nano-agent-enhanced HR-VWI for stroke risk prediction and prevention.

## Data Availability

The data that support the findings of this study are available from the corresponding author,Shuang Xia, upon reasonable request.

## DISCLOSURE PARAGRAPH

1. Guarantor: The scientific guarantor of this publication is Shuang Xia.
2. Conflict of Interest: The authors of this manuscript declare that they have no relationships with any companies whose products or services may be related to the subject matter of the article.
3. Statistics and Biometry: No complex statistical methods were necessary for this paper.
4. Informed Consent: Written informed consent was waived by the Institutional Review Board.
5. Ethical Approval: This animal study was approved by the Institutional Animal Care and Use Committee of Nankai University (No. 2021-SYDWLL-000425). The study was consistent with regulations for the Administration of Affairs Concerning Experimental Animals of China. There are no ethical/legal conflicts involved in the article.
6. Study subjects or cohorts overlap: No study subjects or cohorts have been previously reported.
7. Methodology
  - diagnostic or prognostic study
  - animal experiment

## KEY POINTS

QUESTION: The effectiveness of the Gd-modified macrophage-target gold nanoclusters-based imaging strategy in predicting carotid plaque vulnerability on a medium-sized animal model?

## PERTINENT FINDINGS

IMPLICATIONS FOR PATIENT CARE: The development of a nanoparticle-based imaging strategy that can precisely predict carotid plaque vulnerability in a noninvasive manner could potentially improve patient care by allowing for earlier identification and treatment of high-risk plaques, thereby preventing acute vascular events.

## Abbreviations

AS: List Atherosclerosis

GdMG NCs: Gd-modified macrophage-target gold nanoclusters

HR-VWI: high-resolution vessel wall imaging

PPI: a novel 16-mer peptide targeted scavenger receptor

AI SR-A: scavenger receptors A

TOF-MRA: time-of-flight magnetic resonance angiography

CV: coefficient of variation

## FIGURES AND LEGENTS

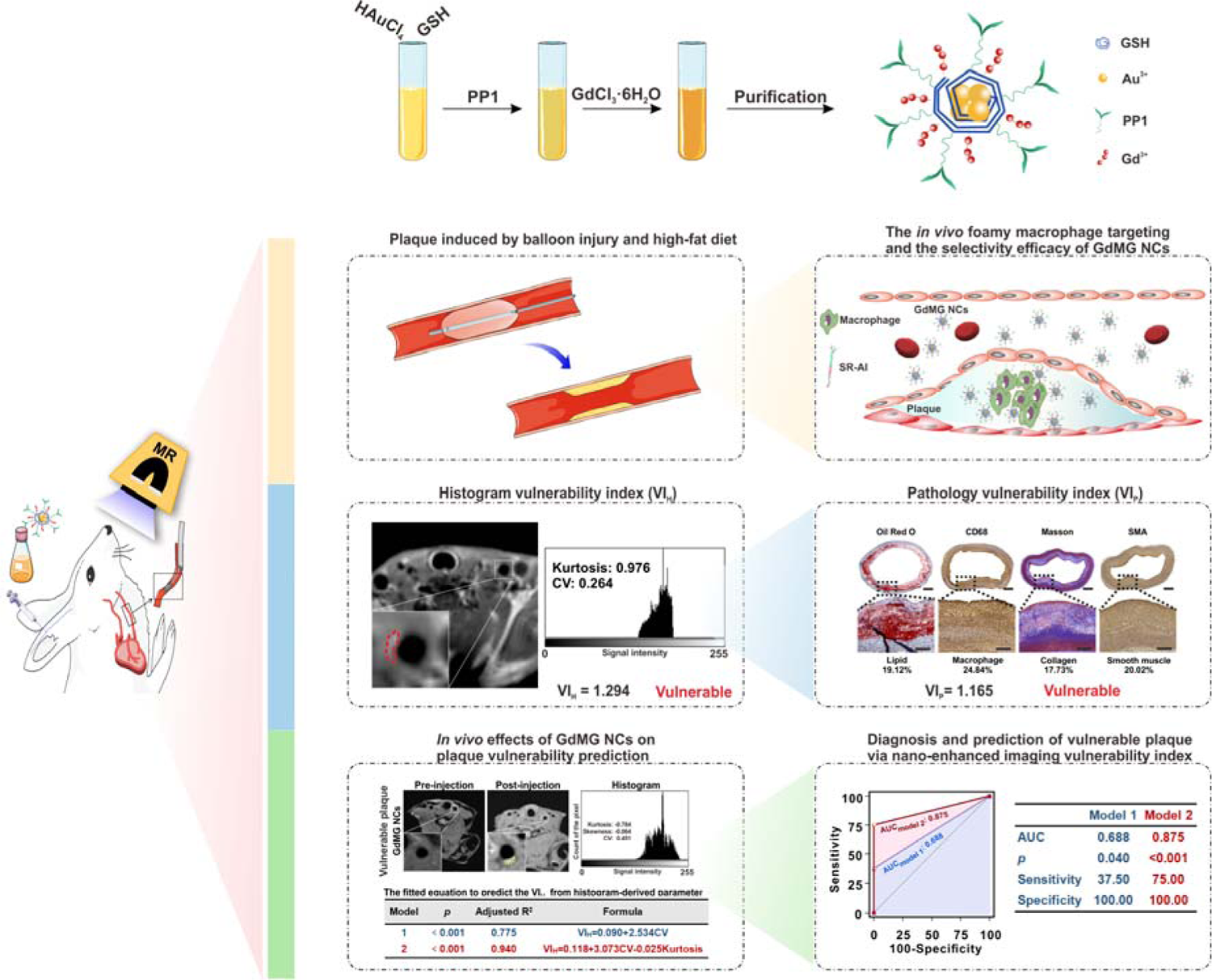
Schematic outlining the synthesis of PP1-Au@GSH@Gd NCs (GdMG NCs) and the strategy for plaque vulnerability prediction.

## REFERENCES

1. Rune I, Rolin B, Lykkesfeldt J. Long-term Western diet fed apolipoprotein E-deficient rats exhibit only modest early atherosclerotic characteristics. Sci Rep 2018;8:5416.

2. Saba L, Moody AR, Saam T. Vessel Wall-Imaging Biomarkers of Carotid Plaque Vulnerability in Stroke Prevention Trials: A viewpoint from The Carotid Imaging Consensus Group. JACC Cardiovasc Imaging 2020;13:2445–2456.

3. Hoshino T, Sissani L, Labreuche J. Prevalence of Systemic Atherosclerosis Burdens and Overlapping Stroke Etiologies and Their Associations With Long-term Vascular Prognosis in Stroke With Intracranial Atherosclerotic Disease. JAMA Neurol 2018;75:203–211.

4. Goddi A, Bortolotto C, Fiorina I. High-frame rate vector flow imaging of the carotid bifurcation. Insights Imaging 2017;8:319–328.

5. Mandell DM, Mossa-Basha M, Qiao Y. Intracranial Vessel Wall MRI: Principles and Expert Consensus Recommendations of the American Society of Neuroradiology. AJNR Am J Neuroradiol 2017;38:218–229.

6. Saba L, Saam T, Jäger HR. Imaging biomarkers of vulnerable carotid plaques for stroke risk prediction and their potential clinical implications. The Lancet Neurology 2019;18:559–572.

7. Zhao X, Hippe DS, Li R. Prevalence and Characteristics of Carotid Artery High-Risk Atherosclerotic Plaques in Chinese Patients With Cerebrovascular Symptoms: A Chinese Atherosclerosis Risk Evaluation II Study. J Am Heart Assoc 2017;6.

8. Kamtchum-Tatuene J, Noubiap JJ, Wilman AH. Prevalence of High-risk Plaques and Risk of Stroke in Patients With Asymptomatic Carotid Stenosis: A Meta-analysis. JAMA Neurol 2020;77:1524–1535.

9. Amirbekian V, Lipinski M, Briley-Saebo K. Detecting and assessing macrophages in vivo to evaluate atherosclerosis noninvasively using molecular MRI. Proc Natl Acad Sci U S A 2007;104:961–966.

10. Li X, Wu M, Li J. Advanced targeted nanomedicines for vulnerable atherosclerosis plaque imaging and their potential clinical implications. Front Pharmacol 2022;13:906512.

11. Xue, Li, Menglin. Ultrasmall bimodal nanomolecules enhanced tumor angiogenesis contrast with endothelial cell targeting and molecular pharmacokinetics. Nanomedicine Nanotechnology Biology & Medicine 2019.

12. Wang J, Wu M, Chang J. Scavenger receptor-AI-targeted ultrasmall gold nanoclusters facilitate in vivo MR and ex vivo fluorescence dual-modality visualization of vulnerable atherosclerotic plaques. Nanomedicine 2019;19:81–94.

13. Wu M, Li X, Guo Q. Magnetic mesoporous silica nanoparticles-aided dual MR/NIRF imaging to identify macrophage enrichment in atherosclerotic plaques. Nanomedicine 2020;32:102330.

14. Obaid DR, Calvert PA, Gopalan D. Atherosclerotic plaque composition and classification identified by coronary computed tomography: assessment of computed tomography-generated plaque maps compared with virtual histology intravascular ultrasound and histology. Circ Cardiovasc Imaging 2013;6:655–664.

15. Visscher M, Moerman AM, Burgers PC. Data Processing Pipeline for Lipid Profiling of Carotid Atherosclerotic Plaque with Mass Spectrometry Imaging. J Am Soc Mass Spectrom 2019;30:1790–1800.

16. Senders ML, Hernot S, Carlucci G. Nanobody-Facilitated Multiparametric PET/MRI Phenotyping of Atherosclerosis. JACC Cardiovasc Imaging 2019;12:2015–2026.

17. Zhang Z, Jiang Y, Zhou Z. Scavenger receptor A1 attenuates aortic dissection via promoting efferocytosis in macrophages. Biochem Pharmacol 2019;168:392–403.

18. Gong Y, Cao C, Guo Y. Quantification of intracranial arterial stenotic degree evaluated by high-resolution vessel wall imaging and time-of-flight MR angiography: reproducibility, and diagnostic agreement with DSA. Eur Radiol 2021.

19. Cao Y, Xiao X, Liu Z. Detecting vulnerable plaque with vulnerability index based on convolutional neural networks. Comput Med Imaging Graph 2020;81:101711.

20. Eslami P, Hartman EMJ, Albaghadai M. Validation of Wall Shear Stress Assessment in Non-invasive Coronary CTA versus Invasive Imaging: A Patient-Specific Computational Study. Ann Biomed Eng 2021;49:1151–1168.

21. Song JW, Pavlou A, Xiao J. Vessel Wall Magnetic Resonance Imaging Biomarkers of Symptomatic Intracranial Atherosclerosis: A Meta-Analysis. Stroke 2021;52:193–202.

22. Shi Z, Li J, Zhao M. Quantitative Histogram Analysis on Intracranial Atherosclerotic Plaques: A High-Resolution Magnetic Resonance Imaging Study. Stroke 2020;51:2161–2169.

23. Michel JB, Lagrange J, Regnault V. Conductance Artery Wall Layers and Their Respective Roles in the Clearance Functions. Arterioscler Thromb Vasc Biol 2022;42:e253–e272.

24. Burtea C, Ballet S, Laurent S. Development of a Magnetic Resonance Imaging Protocol for the Characterization of Atherosclerotic Plaque by Using Vascular Cell Adhesion Molecule-1 and Apoptosis-Targeted Ultrasmall Superparamagnetic Iron Oxide Derivatives. Arteriosclerosis Thrombosis and Vascular Biology 2012;32:E36-+.

25. Trivedi RA, Mallawarachi C, JM UK-I. Identifying inflamed carotid plaques using in vivo USPIO-enhanced MR imaging to label plaque macrophages. Arterioscler Thromb Vasc Biol 2006;26:1601–1606.

26. Howarth SPS, Tang TY, Trivedi R. Utility of USPIO-enhanced MR imaging to identify inflammation and the fibrous cap: A comparison of symptomatic and asymptomatic individuals. Eur J Radiol 2009;70:555–560.

27. Durham AL, Speer MY, Scatena M. Role of smooth muscle cells in vascular calcification: implications in atherosclerosis and arterial stiffness. Cardiovasc Res 2018;114:590–600.

28. Beldman TJ, Senders ML, Alaarg A. Hyaluronan Nanoparticles Selectively Target Plaque-Associated Macrophages and Improve Plaque Stability in Atherosclerosis. Acs Nano 2017;11:5785–5799.

29. Karel M, Hechler B, Kuijpers M. Atherosclerotic plaque injury-mediated murine thrombosis models: advantages and limitations. Platelets 2020;31:439–446.

30. Zhou Y, Zhang NR, Zheng ZN. Regular transient limb ischemia prevents atherosclerosis progression in hypercholesterolemic rabbits. Chin Med J (Engl) 2019;132:1079–1086.

